# Comparison of humoral and cellular responses in kidney transplant recipients receiving BNT162b2 and ChAdOx1 SARS-CoV-2 vaccines

**DOI:** 10.1101/2021.07.09.21260192

**Authors:** Maria Prendecki, Tina Thomson, Candice L. Clarke, Paul Martin, Sarah Gleeson, Rute Cardoso De Aguiar, Helena Edwards, Paige Mortimer, Stacey McIntyre, Shanice Lewis, Jaid Deborah, Alison Cox, Graham Pickard, Liz Lightstone, David Thomas, Stephen P. McAdoo, Peter Kelleher, Michelle Willicombe, in collaboration with the OCTAVE Study Consortium

## Abstract

**Background:** Attenuated immune responses to mRNA SARS-CoV-2 vaccines have been reported in solid organ transplant recipients. Most studies have assessed serological responses alone, and there is limited immunological data on vector-based vaccines in this population. This study compares the immunogenicity of BNT162b2 with ChAdOx1 in a cohort of kidney transplant patients, assessing both serological and cellular responses.

**Methods:** 920 patients were screened for spike protein antibodies (anti-S) following 2 doses of either BNT162b2 (n=490) or ChAdOx1 (n=430). 106 patients underwent assessment with T-cell ELISpot assays. 65 health care workers were used as a control group.

**Results:** Anti-S was detected in 569 (61.8%) patients. Seroconversion rates in infection-naïve patients who received BNT162b2 were higher compared with ChAdOx1, at 269/410 (65.6%) and 156/358 (43.6%) respectively, p<0.0001. Anti-S concentrations were higher following BNT162b, 58(7.1-722) BAU/ml, compared with ChAdOx1, 7.1(7.1-39) BAU/ml, p<0.0001. Calcineurin inhibitor monotherapy, vaccination occurring >1^st^ year post-transplant and receiving BNT162b2 was associated with seroconversion.

Only 28/106 (26.4%) of patients had detectable T-cell responses. There was no difference in detection between infection-naïve patients who received BNT162b2, 7/40 (17.5%), versus ChAdOx1, 2/39 (5.1%), p=0.15. There was also no difference in patients with prior infection who received BNT162b2, 8/11 (72.7%), compared with ChAdOx1, 11/16 (68.8%), p=0.83.

**Conclusions:** Enhanced humoral responses were seen with BNT162b2 compared with ChAdOx1 in kidney transplant patients. T-cell responses to both vaccines were markedly attenuated. Clinical efficacy data is still required but immunogenicity data suggests weakened responses to both vaccines in transplant patients, with ChAdOx1 less immunogenic compared with BNT162b2.

---

Coronavirus disease 2019 (COVID-19) has had a significant impact on immunocompromised patients, including solid organ transplant recipients with significant rates of hospitalisation and severe disease. There have been several reported successful clinical trials of SARS-CoV-2 vaccination, with studies demonstrating the development of robust humoral and cellular immunity and high rates of protection from disease (1, 2). Subsequent ‘real-world’ studies of vaccine efficacy have identified significant protection against both severe and asymptomatic disease in the general population (3, 4) However, immunosuppressed patients, and those with extensive co-morbidity were excluded from vaccine trials.

Published studies have identified that kidney transplant recipients develop impaired humoral responses to other vaccines including influenza, pneumococcal, and hepatitis B (5-7). Despite T-cell directed therapies being the mainstay of immunosuppression regimens for solid organ transplantation, cellular responses to vaccination have not been extensively investigated. Studies of cellular responses to influenza vaccine in solid organ transplant recipients have generated conflicting results (6, 8) These previous studies may also not be applicable to the novel vaccine types such as adenoviral vector and mRNA which are currently utilised for SARS-CoV-2 vaccination.

Evidence is beginning to emerge on the attenuated immunological responses to SARS-CoV-2 mRNA vaccines in solid organ transplant recipients. Serological responses have been reported to occur in only up to 59% of kidney transplant recipients following two doses of mRNA based vaccines (9-13). However, data on responses to adenoviral vector SARS-CoV-2 vaccines in transplant populations are urgently needed.

In this study we describe the serological and T-cell responses to two dose vaccination (with either BNT162b2 mRNA or ChAdOx1 nCoV-19 replication-deficient adenoviral vector vaccines) in a cohort of kidney transplant recipients in order to evaluate the impact of immunosuppression on immunological vaccine responses in this group.

## METHODS

### Study population

Nine-hundred and twenty kidney transplant patients who had received 2 doses of either BNT162b2 or ChAdOx1 were included. All patients were recruited from within Imperial College Renal and Transplant Centre into one of two prospective studies. The first is the OCTAVE study, an Observational Cohort Study of T-cells, Antibodies and Vaccine Efficacy in SARS-CoV-2 in people with chronic diseases and/or secondary immunodeficiency, which is part of the UK COVID-19 Immunity National Core Study Programme. The OCTAVE study was approved by the Health Research Authority, Research Ethics Committee (Reference:21/HRA/0489). The second study is a prospective longitudinal study ‘The effect of COVID-19 on Renal and Immunosuppressed patients’, sponsored by Imperial College London. This study was approved by the Health Research Authority, Research Ethics Committee (Reference: 20/WA/0123).

A subgroup of 106 (11.5%) patients underwent more in-depth immunological analysis of serological and cellular responses to SARS-CoV2 vaccination. A group of 65 healthcare workers (HCW), with a median age of 38 (30-46) years, were used as a comparator. Fifty and 15 HCW received the BNT162b2 and ChAdOx1 vaccines, respectively. The median interval between vaccinations in the HCW was 68 (61-70) days, with a median time to sampling post-vaccination of 28 (21-28) days.

### Serological testing

Serum was tested for antibodies to both the nucleocapsid protein (anti-NP) and spike protein (anti-S). Anti-NP was tested using the Abbott Architect SARS-CoV-2 IgG 2 step chemiluminescent immunoassay (CMIA) according to manufacturer’s instructions. This is a non-quantitative assay and samples were interpreted as positive or negative with a threshold index value of 1.4. The presence of anti-NP was used as a marker of natural infection. For vaccine responses, spike protein antibodies (anti-S IgG) were assessed using the Abbott Architect SARS-CoV-2 IgG Quant II CMIA. Anti-S antibody titres are quantitative with a threshold value of 7.1 BAU/ml for positivity, and an upper level of detection of 5680 BAU/ml.

### T cell ELISpot

SARS-CoV-2 specific T-cell responses were detected using the T-SPOT® Discovery SARS-CoV-2 (Oxford Immunotec) according to the manufacturer’s instructions. In brief, peripheral blood mononuclear cells (PBMCs) were isolated from whole blood samples with the addition of T-Cell Select™ (Oxford Immunotec) where indicated. 250,000 PBMCs were plated into individual wells of a T-SPOT® Discovery SARS-CoV-2 plate. The assay measures immune responses to SARS-CoV-2 spike protein peptide pools (S1 protein and S2 protein), in addition to positive PHA (phytohemagglutinin) and negative controls. Cells were incubated and interferon-γ secreting T cells were detected. Spot forming units (SFU) were detected using an automated plate reader (Autoimmun Diagnostika). Infection-naïve, unvaccinated participants were used to identify a threshold for a positive response using mean +3 standard deviation SFU/10^6^ PBMC, as previously described. This resulted in a cut-off for positivity of 40 SFU/10^6^ PBMC.

### Definition of prior infection

Prior infection was defined serologically or via past PCR positive confirmed infection. The detection of anti-NP on current or historic samples, and the presence of anti-S at baseline (pre-vaccine) or historic samples, was required for the definition of prior infection by serological methods. Serological screening of transplant patients had commenced in our centre in June 2020, and these data was used to aid identification of patients with prior SARS-CoV-2 exposure prior to vaccination. As part of this protocol, patients were initially screened for anti-NP, and those with a subthreshold anti-NP index value (0.25-1.4), underwent confirmatory testing for natural infection by assessing for receptor binding domain (anti-RBD) antibodies. This was performed using an in-house double binding antigen ELISA (Imperial Hybrid DABA; Imperial College London, London, UK), which detects total RBD antibodies(14-17). The HCW had serological analysis at baseline, which was used to define prior infection alone.

### Statistical Analysis

Statistical analysis was conducted using Prism 9.0 (GraphPad Software Inc., San Diego, California). Unless otherwise stated, all data are reported as median with interquartile range (IQR). The Chi-squared test was used for proportional assessments. The Mann-Whitney and Kruskal-Wallis tests were used to assess the difference between 2 or >2 groups, with Dunn’s post-hoc test to compare individual groups. Multivariable analysis was carried out using multiple logistic regression using variables which were found to be significant on univariable analysis.

## RESULTS

Nine-hundred and twenty kidney transplant recipients underwent serological testing for SARS-CoV-2 antibodies at a median time of 31 (27-35) days following their vaccine boost. One-hundred and fifty-two (16.5%) patients had evidence of prior infection. 490 (53.3%) patients received the BNT162b2 vaccine and 430 (46.7%) received the ChAdOx1 vaccine (**Figure 1**). There was no difference in the proportion of patients with prior infection between those receiving BNT162b2, 80 (16.3%), compared with ChAdOX1, 72 (16.7%), p=0.86.

**Figure 1.**
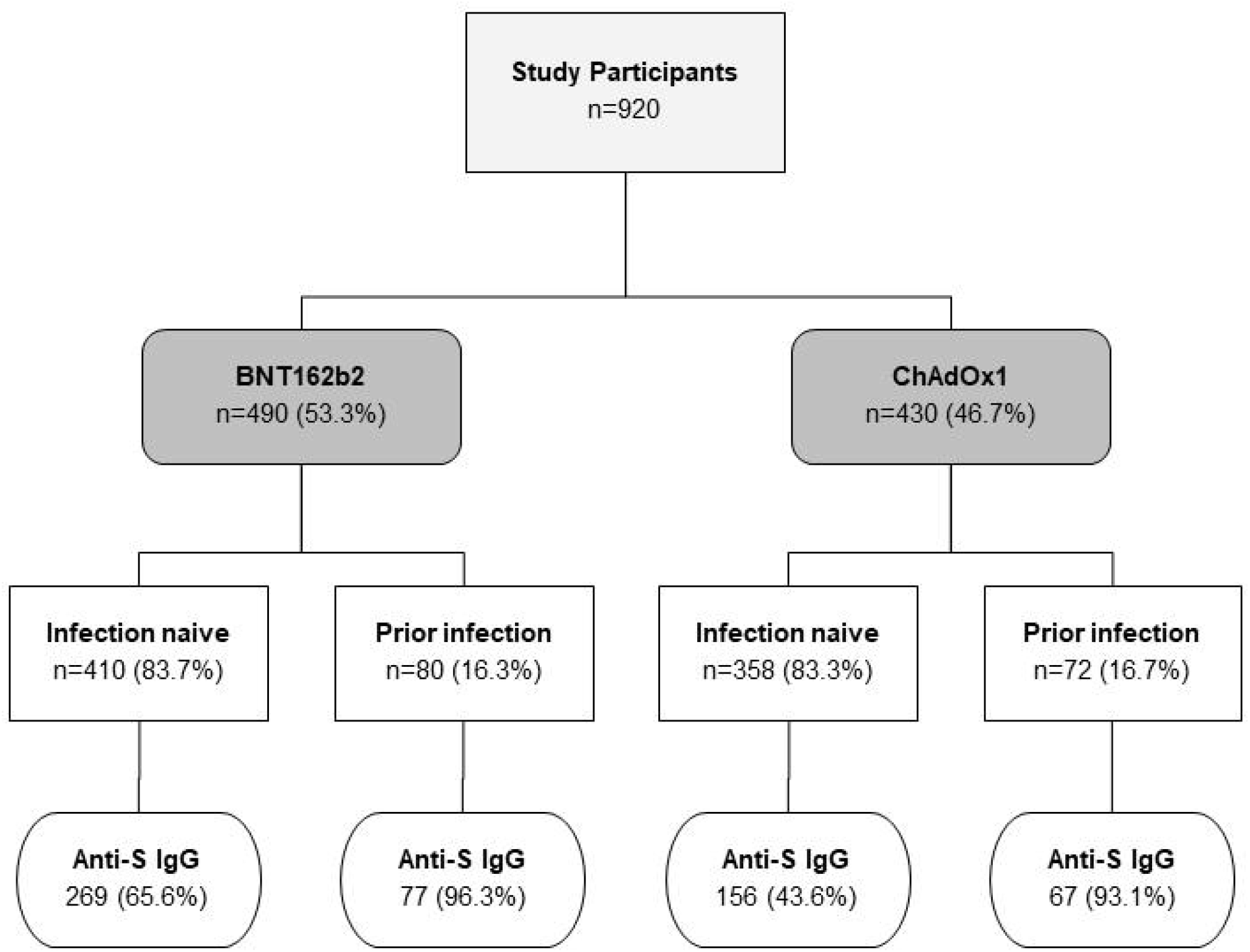
Study flow diagram.

### Serological responses in infection-naïve patients

At a median of 31 (27-35) days post their second-dose vaccine, 425/768 (55.3%) infection-naïve patients had detectable antibodies against the spike protein (anti-S). Patients who received BNT162b2 were more likely to seroconvert compared with patients who received ChAdOx1, with seroconversion rates of 269/410 (65.6%) and 156 (43.6%) respectively, p<0.0001 (**Figure 1**).

On univariable analysis, additional patient characteristics associated with seroconversion in infection-naïve patients included i. receiving tacrolimus monotherapy as maintenance immunosuppression, ii. having received the depleting anti-CD52 antibody, Alemtuzumab, as an immunosuppression induction agent, iii. being non-diabetic and iv. being vaccinated after the 1^st^ year post-transplant **(Table 1)**. On multivariable analysis, receiving tacrolimus monotherapy, OR 5.22 (3.60-7.65), p<0.0001 and being vaccinated with BNT162b2, OR 2.47 (1.79-3.43), p<0.0001, were associated with seroconversion. By contrast, vaccination occurring within the first-year post-transplant, OR 0.28 (0.15-0.55), p=0.0002, and a diagnosis of diabetes, OR 0.65 (0.46-0.92), p=0.015, were associated with a reduced likelihood of seroconversion (*Supplemental Information* **Table S1**).

**Table 1.** **Clinical characteristics associated with seroconversion following SARS-CoV-2 vaccination in infection-naïve kidney transplant recipients**

| Characteristic |  | Failure to seroconvert<br>N=343 (%) | Seroconversion<br>N=425 (%) | p value |
| --- | --- | --- | --- | --- |
| **Gender** | Male | 228 (66.3) | 283 (65.7) | 0.86 |
|  | Female | 116 (33.7) | 148 (34.3) |  |
| **Age** | Years (Median) | 60 (49-67) | 59 (48-67) | 0.30 |
| **Ethnicity** | Caucasian | 164 (47.8) | 200 (47.1) | 0.14 |
|  | Black | 31 (9.0) | 26 (6.1) |  |
|  | Indoasian | 94 (27.4) | 143 (33.6) |  |
|  | Other | 54 (15.7) | 56 (13.2) |  |
| **Cause of ESKD** | Polycystic kidney disease | 40 (11.7) | 51 (12.0) | 0.74 |
|  | Glomerulonephritis | 106 (30.9) | 123 (28.9) |  |
|  | Diabetic nephropathy | 69 (20.1) | 75 (17.6) |  |
|  | Urological | 27 (7.9) | 39 (9.2) |  |
|  | Unknown | 66 (19.2) | 98 (23.1) |  |
|  | Other | 35 (10.2) | 39 (9.2) |  |
| **Vaccinated ≤1 year post transplant** | Yes | 42 (12.2) | 17 (4.0) | <0.0001 |
|  | No | 301 (87.8) | 408 (96.0) |  |
| **Time vaccinated post transplant** | Years (Median) | 6.8 (2.4-13.6) | 6.7 (3.2-11.9) | 0.78 |
| **Immunosuppression at diagnosis** | CNI monotherapy* | 97 (28.3) | 284 (66.8) | <0.0001 |
|  | CNI/anti-proliferative | 123 (35.9) | 74 (17.4) |  |
|  | CNI/steroids | 21 (6.1) | 26 (6.1) |  |
|  | CNI/anti-proliferatives/steroids | 95 (27.7) | 36 (8.5) |  |
|  | Anti-proliferatives/steroids | 3 (0.9) | 1 (0.2) |  |
|  | Other | 4 (1.2) | 4 (0.9) |  |
| **Induction immunotherapy** | Alemtuzumab*<br>IL2 receptor blocker<br>None/Unknown/Other | 194 (56.6)<br>60 (17.5)<br>89 (25.9) | 333 (78.4)<br>23 (5.4)<br>69 (16.2) | **<0.0001** |
| **Transplant number** | 1<sup>st</sup><br>≥2<sup>nd</sup> | 292 (85.1)<br>51 (14.9) | 380 (89.4)<br>45 (10.6) | 0.07 |
| **Diabetes** | No<br>Yes | 210 (61.2)<br>133 (38.8) | 294 (69.2)<br>131 (30.8) | **0.021** |
| **Vaccine type** | BNT1262b2<br>ChAdOx1 | 141 (41.1)<br>202 (58.9) | 269 (63.3)<br>156 (36.7) | **<0.0001** |
| **Time between vaccinations** | Days (median) | 74 (66-78) | 75 (68-77) | 0.76 |
| **Time of serological test post-boost** | Days (median) | 31 (27-35) | 31 (27-35) | 0.49 |

CNI=calcineurin inhibitors

Serum concentrations of anti-S were significantly higher in patients who received BNT162b compared with ChAdOx1, with median anti-S concentrations of 58 (7.1-722) BAU/ml and 7.1 (7.1-39) BAU/ml respectively, p<0.0001 (**Figure 2a**). Details of the correlation between individual clinical characteristics and anti-S levels may be found in the Supplemental Information (*Supplemental Information*, **Figure S1**).

**Figure 2.**
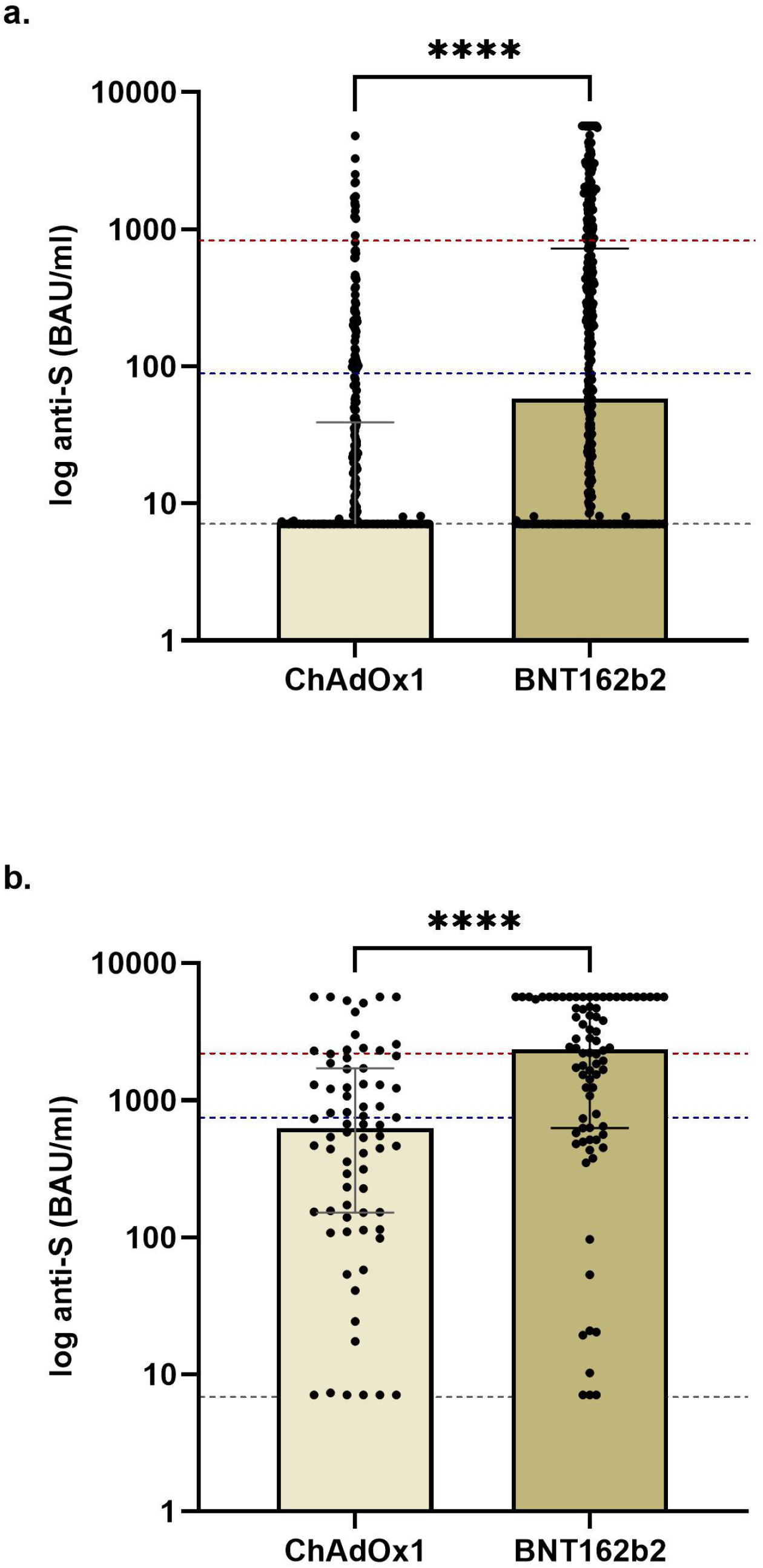
Comparison of anti-S concentrations in patients receiving BNT162b2 versus ChAdOx1 by prior exposure. a. Infection naïve kidney transplant patients who received BNT162b2 had significantly higher anti-S concentrations, 58 (7.1-722) BAU/ml compared with ChAdOx1, 7.1 (7.1-39) BAU/ml, p<0.0001. The red and blue dotted lines represent the median anti-S in infection-naïve HCW receiving BNT162b2 and ChAdOx1 respectively. b. Patients with prior infection who received BNT162b2 had significantly higher anti-S concentrations, 2350 (628-5680) BAU/ml compared with ChAdOx1, 622 (151-1706) BAU/ml, p<0.0001. The red and blue dotted lines represent the median anti-S in previously exposed HCW receiving BNT162b2 and ChAdOx1 respectively.

### Serological responses in patients with prior infection

Patients with prior infection were highly likely to be seropositive, with only 8/152 (5.3%) patients remaining seronegative post-vaccination; 5/72 (6.9%) who received ChAdOx1 and 3/80 (3.7%) who received BNT162b2, p=0.38. Of the 8 patients, 3 had prior infection diagnosed by PCR testing, 4 patients had current anti-NP positivity and 1 patient had historic anti-RBD positivity. Patients who received ChAdOx1 had a median anti-S of 622 (151-1706) BAU/ml, which was significantly less than patients who had received BNT162b2, who had a median anti-S concentration of 2350 (628-5680) BAU/ml, p<0.0001 (**Figure 2b**). Adjusting for vaccine type, there was no difference in post-vaccination anti-S in patients with prior infection diagnosed by PCR compared with serology, with median anti-S concentrations of 324 (111-844) and 758 (172-1985) BAU/ml respectively, in patients receiving ChAdOx1, p=0.59; and 1659 (546-3684) and 2752 (641-5680) BAU/ml respectively, in patients receiving BNT162b2, p=0.76 (*Supplemental Information* **Figure S2)**. There was no difference in post-vaccination anti-S in patients who were anti-NP positive compared with anti-NP negative at the time of testing, with median concentrations of 503 (37-2094) and 701 (153-1694) BAU/ml respectively, in patients receiving ChAdOx1, p=0.91; and 2512 (499-5680) and 2350 (780-5022) BAU/ml respectively, in patients receiving BNT162b2, p=0.99 (*Supplemental Information* **Figure S2)**.

### Cellular and humoral responses in infection-naïve patients

A more in-depth analysis was performed on a subgroup of 106 patients. They underwent paired assessment of T-cell and serological responses at a median time of 31 (29-34) days post-vaccination.

A comparison of clinical characteristics between the sub- and main groups can be found in the *Supplemental Information*, **Table S2**. Fifty-one of 106 (48.1%) patients received BNT162b2 and 55 (51.9%) received ChAdOx1. Forty of 51 (78.4%) patients who received BNT162b2 and 39/55 (70.9%) of those who received the ChAdOx1 vaccine were infection-naïve, p=0.38. A comparison of clinical characteristics of infection-naïve patients by vaccine administered can be found in the *Supplemental Information*, **Table S3**.

Only 9/79 (11.4%) infection-naïve patients had detectable T-cell responses post-vaccination. There was no proportional difference in ELISpot positivity in infection-naïve patients who received BNT162b2, 7/40 (17.5%), compared with ChAdOx1, 2/39 (5.1%), p=0.15. Quantifying cellular responses, infection-naïve patients who received BNT162b2 had a greater T-cell responses compared with patients who received ChAdOx1, with 14 (4-32) SFU/10^6^ PBMCs and 4 (0-12) SFU/10^6^ PBMCs respectively, p=0.019. Notably, T-cell responses in infection-naïve transplant patients were far inferior to those of infection-naïve healthcare workers. Infection-naïve HCW receiving BNT162b2 or ChAdOx1 had a median 63 (21-132) SFU/106 PBMCs, p=0.0003 and 68 (30-162) SFU/106 PBMCs, p<0.0001, respectively (**Figure 3a**).

**Figure 3.**
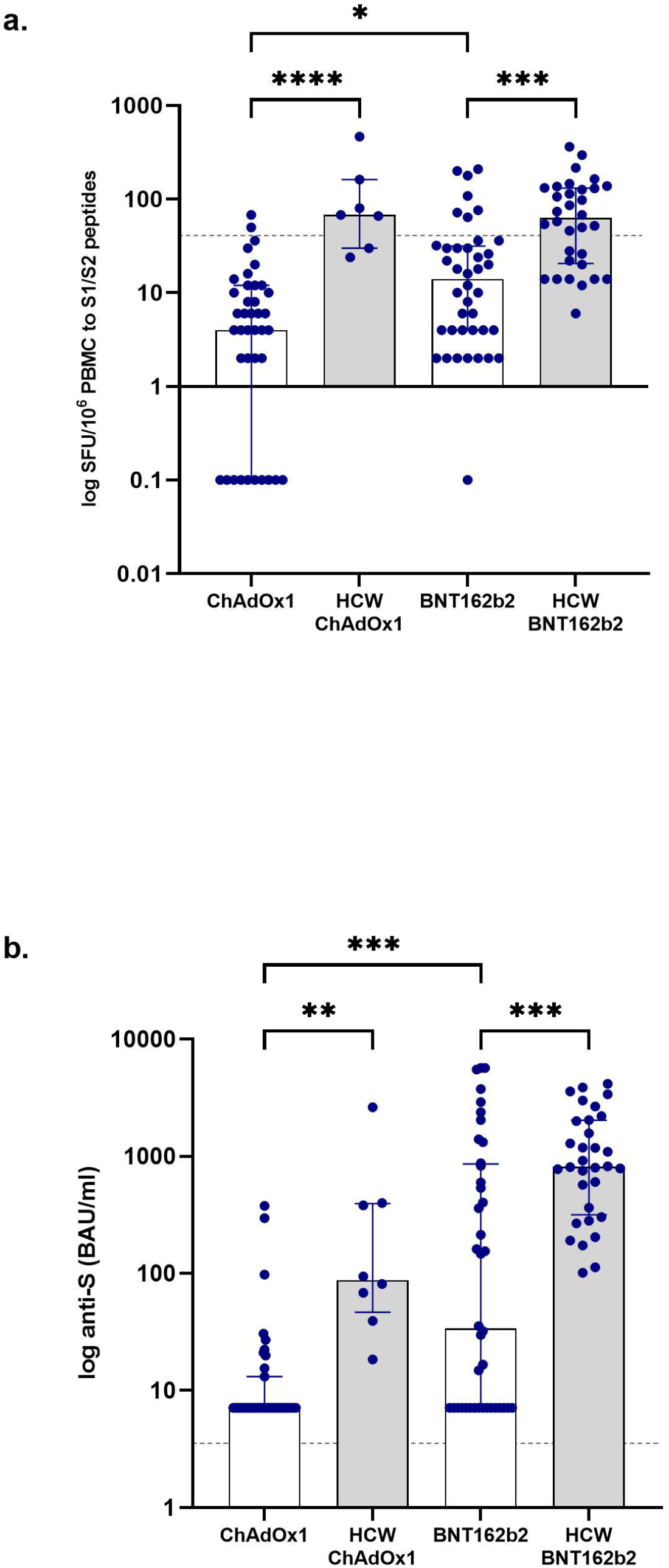
Comparison of post-vaccination T-cell and serological responses in infection naïve patients and healthcare workers. a. Infection naïve patients who received BNT162b2 had a greater T-cell response compared with patients who received ChAdOx1, with 14 (4-32) SFU/10^6^ PBMCs and 4 (0-12) SFU/10^6^ PBMCs respectively, p=0.019. Infection naïve HCW receiving BNT162b2 and ChAdOx1 had significantly greater responses compared with patients having the corresponding vaccine, with a median 63 (21-132) SFU/10^6^ PBMCs, p=0.0003 and 68 (30-162) SFU/10^6^ PBMCs, p<0.0001 respectively. b. Anti-S concentrations in patients who received BNT162b2 were higher compared with those patients who had received ChAdOx1, with anti-S concentrations of 34 (7.1-861) BAU/ml and 7.1 (7.1-13) BAU/ml respectively, p=0.0005. These anti-S concentrations in patients were significantly lower than infection naïve HCW who received the corresponding vaccine, with HCW receiving BNT162b2 and ChAdOx1 having a median anti-S of 815 (318-2033), p=0.0003, and 88 (47-395) BAU/ml, p=0.01, respectively.

Seroconversion rates in infection-naïve patients following vaccination with BNT162b2 were significantly higher compared with vaccination with ChAdOx1, with 24/40 (60.0%) and 10/39 (25.6%) patients seroconverting respectively, p=0.002. In keeping with this, anti-S concentrations in infection-naïve patients who received BNT162b2 were higher compared with those patients who had received ChAdOx1, with anti-S concentrations of 34 (7.1-861) BAU/ml and 7.1 (7.1-13) BAU/ml respectively, p=0.0005 (**Figure 3b**). Serum concentrations of anti-S in infection-naïve patients were significantly lower than infection-naïve HCW who received the corresponding vaccine, with HCW receiving BNT162b2 having a median anti-S of 815 (318-2033), p=0.0003, and those receiving ChAdOx1 a median anti-S of 88 (47-395) BAU/ml, p=0.01 (**Figure 3b**).

### Cellular and humoral responses in patients with prior infection

Overall, 19/27 (70.4%) of patients with prior exposure had detectable T-cell responses post-vaccination. There was no proportional difference in ELISpot positivity in patients with prior infection who received BNT162b2, 8/11 (72.7%), compared with ChAdOx1, 11/16 (68.8%), p=0.83. Quantification of cellular responses found no differences in median SFU/10^6^ PBMCs in previously exposed patients who received BNT162b2 compared with ChAdOx1, with 160 (20-422) SFU/10^6^ PBMCs and 95 (8-144) SFU/10^6^ PBMCs respectively, p=0.42 (**Figure 4a**). In addition, there were no significant differences between T-cell responses in previously exposed patients and HCW receiving the corresponding vaccine, with previously exposed HCW receiving BNT162b2 or ChAdOx1 having a median 246 (131-332) SFU/10^6^ PBMCs, p=0.63 and 114 (74-216) SFU/10^6^ PBMCs, p=0.85 respectively (**Figure 4a**).

**Figure 4.**
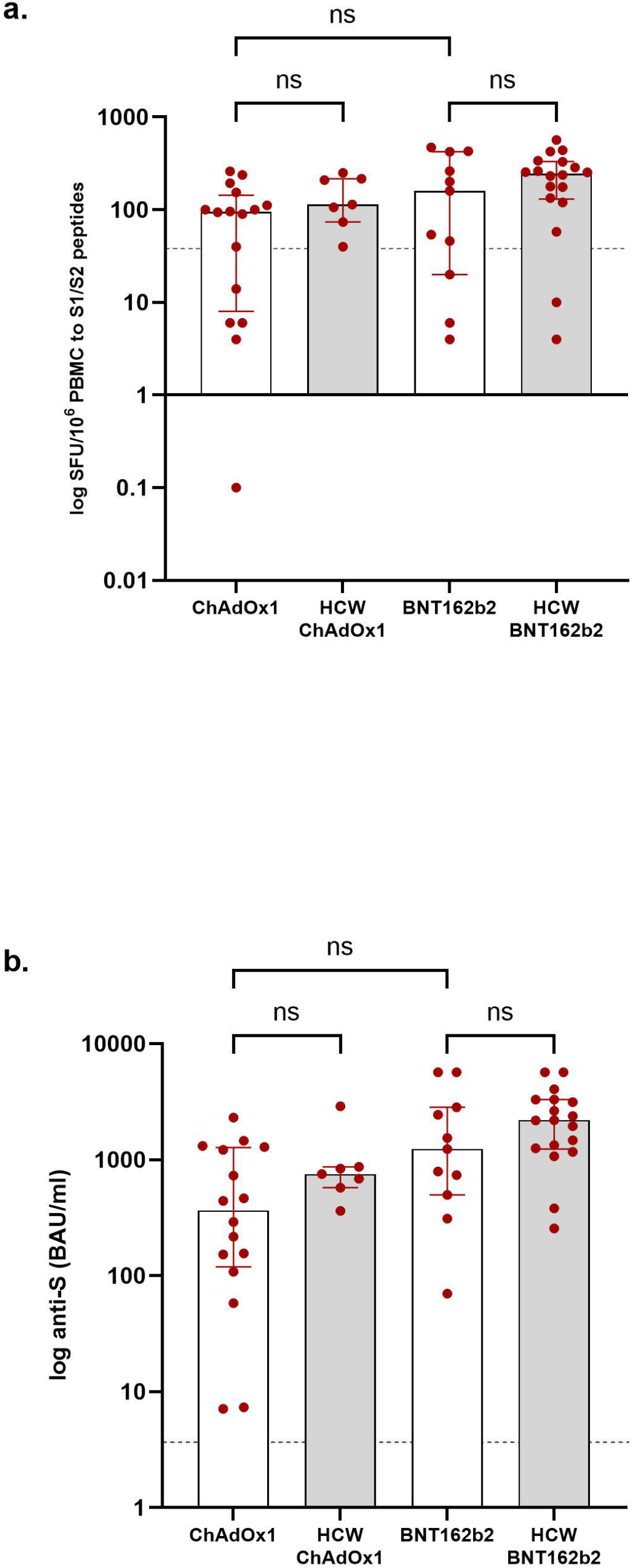
Comparison of post-vaccination T-cell and serological responses in patients and healthcare workers with prior exposure to SARS-CoV-2. **a**. There was no difference in median SFU/10^6^ PBMCs in previously exposed patients who received BNT162b2 compared with ChAdOX1, with 160 (20-422) SFU/10^6^ PBMCs and 95 (8-144) SFU/10^6^ PBMCs respectively, p=0.42. There were no differences between T-cell responses in previously exposed patients and HCW receiving the corresponding vaccine, with HCW receiving BNT162b2 and ChAdOx1 having a median 246 (131-332) SFU/10^6^ PBMCs, p=0.63 and 114 (74-216) SFU/10^6^ PBMCs, p=0.85 respectively **b**. There was no difference in anti-S concentrations in patients with prior infection who received BNT162b2, 1238 (497-2829) BAU/ml, compared with ChAdOx1, 366 (119-1273) BAU/ml, p=0.07. There were also no differences between patients and HCW with prior exposure who received BNT162b2, median anti-S 2189 (1236-3303) BAU/ml), p=0.72, or HCW who received ChAdOx1, median anti-S 753 (574-867) BAU/ml, p=0.91

Twenty-six of 27 (96.3%) patients with prior evidence of infection had detectable anti-S post-vaccination. There was no difference in anti-S concentrations in patients with prior infection who received BNT162b2, 1238 (497-2829) BAU/ml, compared with ChAdOx1, 366 (119-1273) BAU/ml, p=0.07. There were also no differences between patients and HCW with prior exposure who received BNT162b2, median anti-S 2189 (1236-3303) BAU/ml, p=0.72, or ChAdOx1, median anti-S 753 (574-867) BAU/ml, p=0.91 (**Figure 4b**).

Within the 106 patients assessed, a positive correlation between serological (anti-S) and cellular responses (SFU/10^6^ PBMCs) was seen, r=0.53, p<0.0001 (*Supplemental Information*, **Figure S3**). Overall, 45/106 (42.5%) of patients within the subgroup had no detectable immunological response, this represented 44/79 (55.7%) of infection-naïve patients and 1/27 (3.7%) of patients with prior exposure. On univariable analysis, vaccination after the first-year post-transplant, Alemtuzumab induction, CNI monotherapy, BNT162b2 and a shorter time to immunological testing post-vaccination were associated with greater chance of a detectable immune response (*Supplemental Information*, **Table S3**). On multivariable analysis, receiving CNI monotherapy was associated with greater chance of an immune response, OR 16.5 (4.7-58.0), p<0.0001; whilst being vaccinated within the first-year post-transplant was associated with no detectable immune response by either measure, OR 0.14 (0.04-0.57), p=0.006 (*Supplemental Information*, **Table S4**).

## DISCUSSION

Whilst it is generally acknowledged that vaccines utilising mRNA platforms generate greater antibody responses compared with vector based vaccines, the latter have been considered superior in their ability to produce robust cellular responses(18). However, to our knowledge no direct comparison has been reported in solid organ transplant recipients who are predominantly maintained on T-cell directed immunosuppression. In this study comparing the immunogenicity of BNT162b2 versus ChAdOx1 vaccines in kidney transplant recipients, we have shown that BNT162b2 induces greater spike protein antibody responses compared with ChAdOx1 in infection-naïve patients. The superior cellular responses we observed with BNT162b2 in the infection naïve patients’ needs careful interpretation given the clinical differences between the cohorts, in addition, when the magnitude of T-cell responses are so poor, meaningful comparison is difficult. However, after adjusting for the clinical factors associated with non-response within the ChAdOx1 group, at most, comparable T-cell responses remain. Hence, the benefit of robust cellular responses to ChAdOx1 were not seen in this transplant population.

Data on mRNA vaccines in transplant patients has demonstrated the contribution of patient related factors in serological response, with older age and immunosuppression type dominating the risk associated with non-response(11, 12, 19). In this study, the reported seroconversion rate of 269/410 (65.6%) in the infection-naïve patients receiving BNT162b2 is numerically higher than that reported from other series assessing responses to mRNA-based vaccines(10-13, 19-21). This may reflect the common use of calcineurin inhibitor monotherapy, in the absence of anti-proliferative agents or corticosteroids in this cohort. In regards to serological responses to vector-based vaccines, only data from a single study of 12 transplant patients is currently available, which reports seroconversion of 2/12 (17%) patients who had received the Ad26.COV2.S vaccine(22).

Opposing the observation of enhanced serological responses in our cohort, immunogenicity studies of mRNA vaccines in transplant recipients assessing cellular responses, have reported better T-cell responses compared with the data we present. Up to 57% of transplant recipients have been shown to develop detectable T-cell responses following vaccination with either mRNA-1273 or BNT162b2(10, 12, 23). These reported differences may relate to the detection methods, or the patient population studied. Alemtuzumab, an anti-CD52 monoclonal antibody was the most commonly used induction agent in the participants of this study, and its use can be associated for prolonged lymphopenia. The poor responses to PHA seen in 7/106 (6.7%) patients in our study highlights the weakened functionality of T-cells in some transplant patients. From a methodological perspective, there may be differences in the peptide pools used in the ELISpot assays, or the SFU threshold we used to define T-cell positivity. The positive threshold we used was set using healthy controls, which may lack the sensitivity to detect T-cell responses in immunosuppressed patients. The peptides we used exclude those with homologies to common human coronaviruses, to minimise cross-reactivity when assessing vaccine efficacy. In addition, the ELISpot assay uses IFN-γ as the sole read out for T-cell reactivity, underestimating T-cell responses overall(24, 25). These factors may contribute to the dissociation we found between serological and cellular responses, as a significant number of patients had detectable antibodies in the absence of T-cell responses.

This study has several limitations, some of which have already been outlined above. In addition, it is important we highlight that the subgroup of patients who underwent T-cell analysis differ from the main cohort. They were younger, more likely to have been vaccinated in the first-year post-transplant and received their 2^nd^ dose of vaccine at a shorter interval compared with the main cohort, albeit at a median time of >8 weeks. This reflected clinical prioritisation for newly transplanted patients, with a high exposure risk, to be vaccinated first. Also, the healthy control reference group, is not a matched control group, with healthcare workers being significantly younger than our patient cohort, which independently could result in better immunological responses independent of immunosuppression.

Notwithstanding these limitations, this study has demonstrated markedly diminished humoral and cellular immune responses to both the BNT162b2 and ChAdOx1 vaccines in kidney transplant patients. No enhanced cellular responses were seen with ChAdOx1, however our results do demonstrate inferior serological responses in patient’s receiving ChAdOx1 compared with BNT162b2. Although it is acknowledged that immunogenicity of vaccines does not necessarily equate to clinical efficacy in this immunosuppressed or other populations, emerging data of SARS-CoV-2 related deaths in transplant recipients, may suggest a correlation between absence of detectable immunological response and efficacy(26, 27). The data we report supports the need to vaccinate people prior to starting immunosuppression if possible. The planning of intervention studies to optimise vaccine platform and dosing are urgently required, with some early encouraging responses to 3^rd^ vaccine doses in transplant recipients(28, 29). Reduction in immunosuppression to enhance responses is not endorsed in transplant recipients, who may risk allograft rejection and premature failure, in so doing. Therefore, in the interim, strategic planning to protect this vulnerable population is required until more data are available. This may include but is not limited to, educating patients to maintain social distancing rules and immunising household members including prioritisation of children ≥12 years.

## Supporting information

Strobe

## Data Availability

The data is not openly available on an external link. Requests for data should be made to the corresponding author.

## ACKNOWLEDGEMENTS

The OCTAVE trial, which is part of the COVID-19 Immunity National Core Study Programme, was sponsored by the University of Birmingham and funded by a grant from UK Research and Innovation (UKRI) administered by the Medical Research Council (grant reference number MC_PC_20031). It has been designated an Urgent Public Health (UPH) study by the National Institute of Health Research.

This research is also supported by the National Institute for Health Research (NIHR) Biomedical Research Centre based at Imperial College Healthcare NHS Trust and Imperial College London. The authors would like to thank the West London Kidney Patient Association, all the patients and staff at ICHNT (The Imperial COVID vaccine group and dialysis staff, and staff within the North West London Pathology laboratories). The authors are also grateful for the support from Hari and Rachna Murgai, The Nan Diamond Fund and the Auchi Charitable Foundation. MP is supported by an NIHR clinical lectureship.

