## Supplemental Information for "Comparison of humoral and cellular responses in kidney transplant recipients receiving BNT162b2 and ChAdOx1 SARS-CoV-2 vaccines"

### **Table S1. Multivariable analysis of clinical characteristics associated with seroconversion following SARS-CoV-2 vaccination in kidney transplant recipients.**

| **Variable** | **Reference Group** | **Univariable** | | **Multivariable** | |
| --- | --- | --- | --- | --- | --- |
|  |  | **OR (95% CI)** | **p value** | **OR (95% CI)** | **p value** |
| **Increasing age** |  | 0.99 (0.98-1.01) | 0.07 | 0.98 (0.97-1.00) | **0.065** |
| **Time of vaccine post-transplant** | <1 year | 0.30 (0.17-0.53) | <0.0001 | 0.28 (0.15-0.55) | **0.0002** |
| **Number of transplants** | ≥2 | 0.68 (0.44-1.04) | 0.08 | - | - |
| **Transplant induction agent** | Alemtuzumab | 2.78 (2.03-3.82) | <0.0001 | - | - |
| **Maintenance Immunosuppression** | CNI monotherapy | 5.11 (3.75-6..96) | <0.0001 | 5.22 (3.60-7.65) | **<0.0001** |
| **Vaccine** | BNT1262b2 | 2.47 (1.85-3.31) | <0.0001 | 2.47 (1.79-3.43) | **<0.0001** |
| **Diabetes** | Yes | 0.70 (0.52-0.94) | 0.021 | 0.65 (0.46-0.92) | **0.015** |

CNI=calcineurin inhibitors

### **Figure S1. Correlation between anti-S concentrations and clinical characteristics in infection naïve patients.**

a. Kidney transplant patients who were receiving calcineurin inhibitor (CNI) monotherapy had significantly higher anti-S concentrations, 75 (7.1-646) BAU/ml, compared with patients receiving CNI in combination with anti-proliferative agents (mycophenolate mofetil or azathioprine), with or without corticosteroids, 7.1 (7.1-28) BAU/ml, p<0.0001
b. Comparing patients receiving combination therapy alone, there was no difference between those who had received Alemtuzumab induction, 7.1 (7.1-67) BAU/ml compared with IL-2 receptor antibodies, 7.1 (7.1-7.12) BAU/ml, p=0.06. Comparing patients who received Alemtuzumab as induction alone, those who are maintained on CNI monotherapy, 73 (7.1-656) BAU/ml, had significantly higher anti-S than those on combination therapy, p<0.0001.
c. There was no difference in anti-S between females, 17 (7.1-264) BAU/ml compared with males, 11 (7.1-223) BAU/ml, p=0.76
d. There was no correlation between cause of end stage kidney disease (ESKD) and anti-S concentrations. Patients with ESKD secondary to diabetes had a median anti-S concentration of 9 (7.1-174) BAU/ml, polycystic kidney disease (APKD), 18 (7.1-415) BAU/ml, Glomerulonephritis (GN), 9 (7.1-238), Urological causes of ESKD, 14 (7.1-205) BAU/ml, and unknown causes of ESKD, 15 (7.1-221) BAU/ml, p=0.83
e. There was no correlation between ethnicity and anti-S concentrations. Patients from Indoasian, Black and White backgrounds having a median anti-S concentration of 19 (7.1-272), 7.1 (7.1-51) and 13 (7.1-270) respectively, p=0.13.
f. Patients who were within their first-year post-transplant when vaccinated had significantly lower anti-S, 7.1 (7.1-21) BAU/ml compared with those patients who were vaccinated after the 1^st^ year, 17 (7.1-271) BAU/ml, p<0.0001.
g. A significant inverse correlation was seen between anti-S response and age in patients who received BNT162b2, r=-0.14, p=0.0038 (i), but not in patients who received ChAdOx1, r=-0.08, p=0.13 (ii).

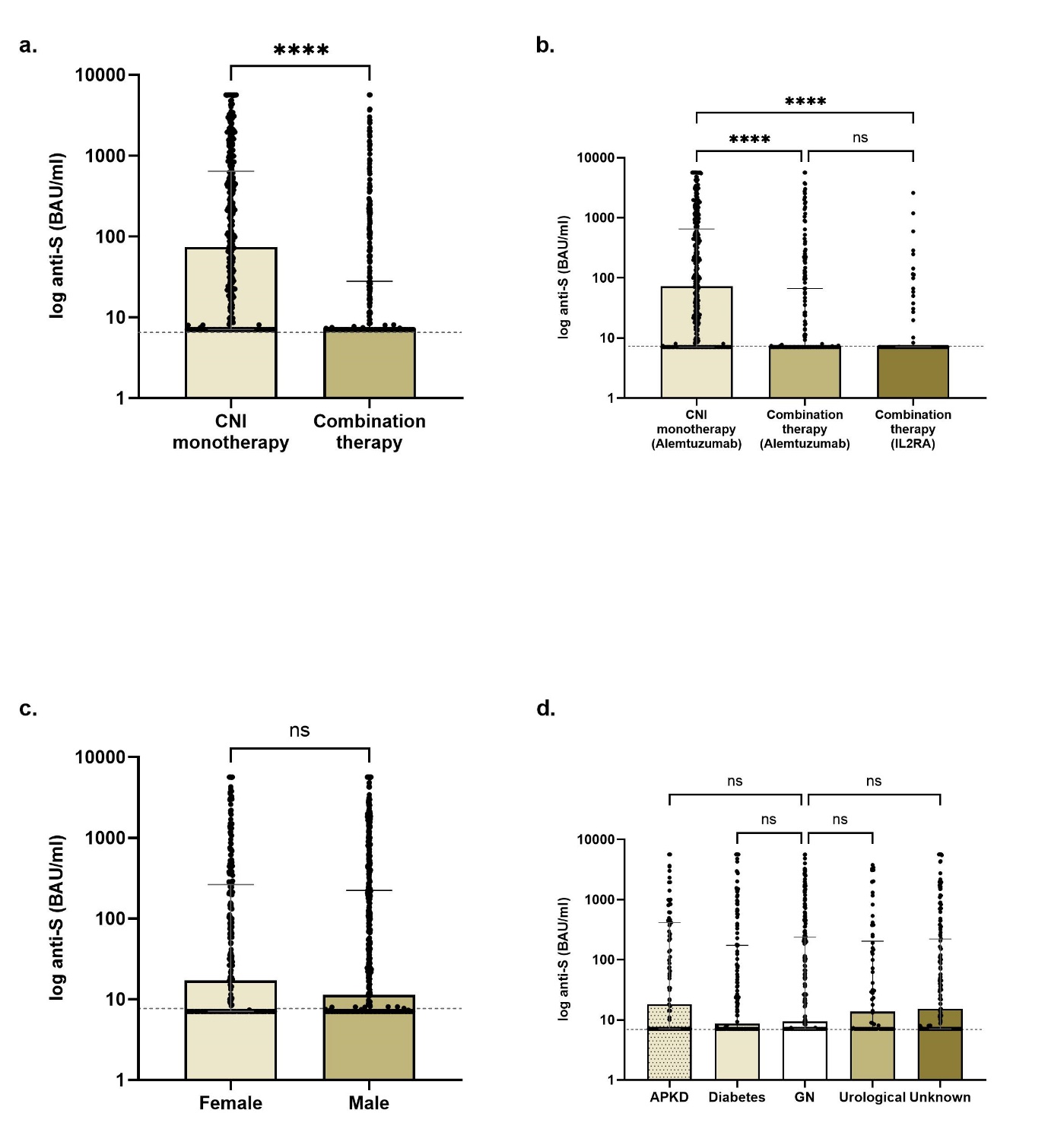

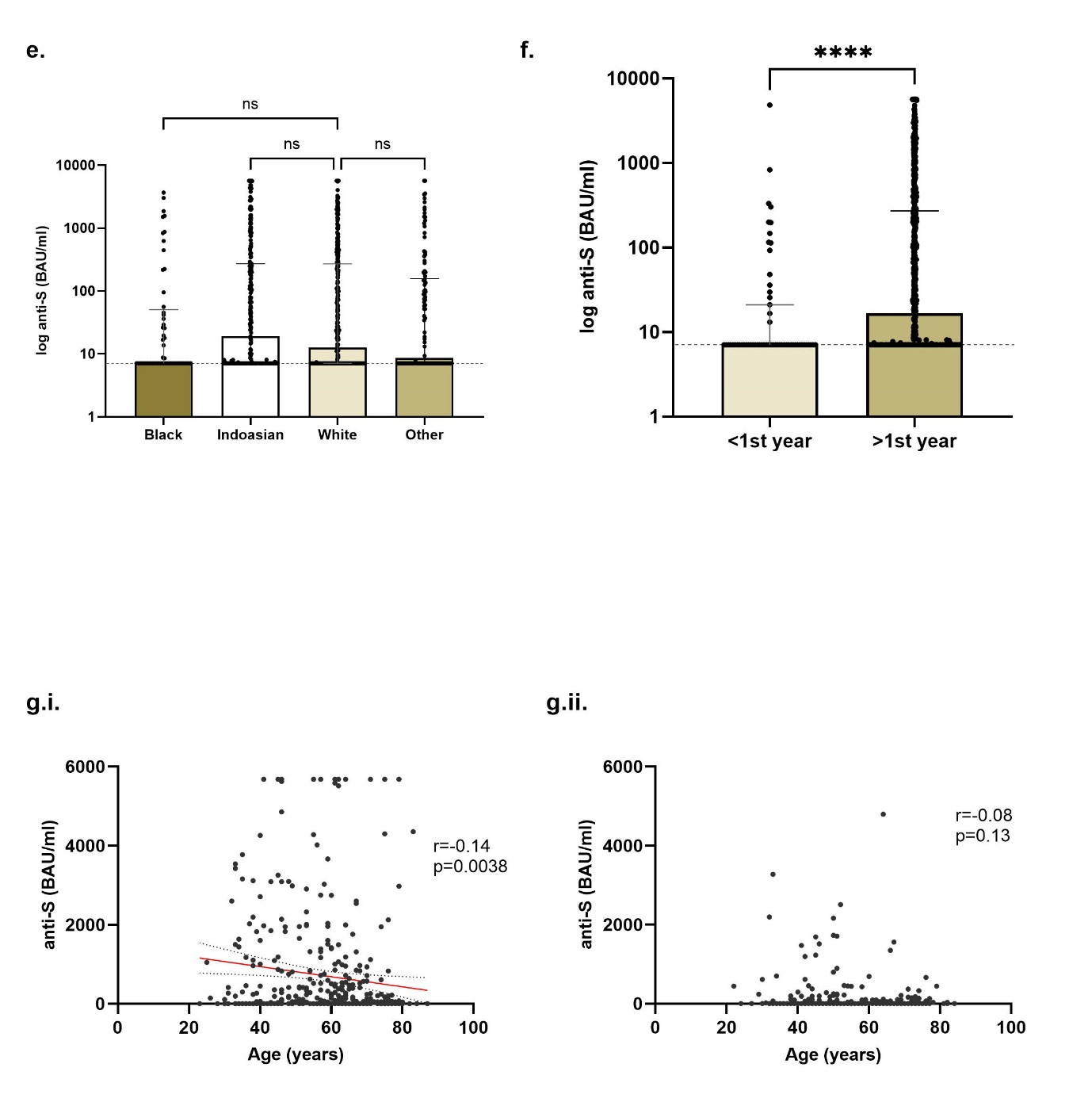

### **Figure S2. Correlation between anti-S concentrations and diagnostic characteristics in patients with prior infection.**

a. Anti-S concentrations in patients who were diagnosed by PCR compared with serology were 324 (111-844) and 758 (172-1985) BAU/ml respectively, p=0.59, in patients receiving ChAdOx1; and 1659 (546-3684) and 2752 (641-5680) BAU/ml respectively, p=0.76, in patients receiving BNT162b2.
b. Anti-S concentrations in patients who were anti-NP positive compared with negative at the time of testing, were 503 (37-2094) and 701 (153-1694) BAU/ml respectively, p=0.91, in patients receiving ChAdOx1; and 2512 (499-5680) and 2350 (780-5022) BAU/ml respectively, p=0.99, in patients receiving BNT162b2

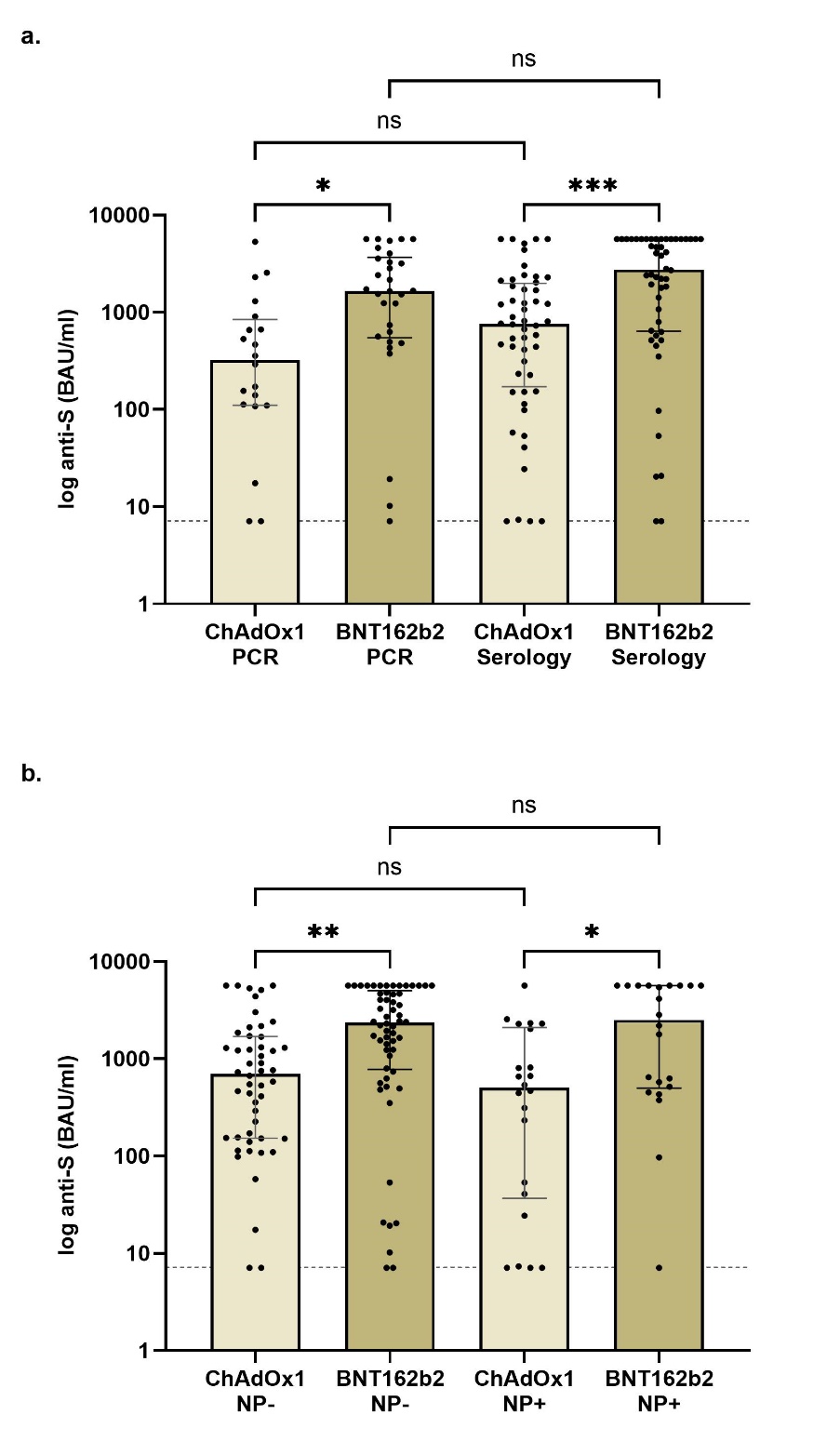

### **Table S2. Comparison of the clinical characteristics of the subgroup of patients who had cellular responses assessed compared with the main cohort.**

| Characteristic | | Main Cohort  N=920 (%) | Subgroup  N=82 (%) | p value |
| --- | --- | --- | --- | --- |
| Gender | Male  Female | 604 (65.7)  316 (34.3) | 54 (65.9)  28 (34.1) | 0.97 |
| Age | Years (Median) | 59 (48-67) | 52 (42-62) | **0.0003** |
| Ethnicity | Caucasian*  Black  Indoasian  Other | 404 (43.9)  84 (9.1)  301 (32.7)  131 (14.2) | 30 (36.6)  12 (14.6)  26 (31.7)  14 (17.1) | 0.20 |
| Cause of ESKD | Polycystic kidney disease  Glomerulonephritis*  Diabetic nephropathy  Urological  Unknown  Other | 107 (11.6)  272 (29.6)  171 (18.6)  72 (7.8)  208 (22.6)  90 (9.8) | 8 (9.8)  26 (31.7)  12 (14.6)  7 (8.5)  21 (25.6)  8 (9.8) | 0.68 |
| Vaccinated ≤1 year post transplant | Yes  No | 92 (10.0)  828 (90.0) | 43 (52.4)  39 (47.6) | **<0.0001** |
| Time vaccinated post transplant | Years (Median) | 6.6 (6.1-7.3) | 0.7 (0.4-7.7) | **<0.0001** |
| Immunosuppression at diagnosis | CNI monotherapy*  CNI/anti-proliferative  CNI/steroids  CNI/anti-proliferatives/steroids  Anti-proliferatives/steroids  Other | 458 (49.8)  240 (26.1)  56 (6.1)  149 (16.2)  4 (0.4)  13 (1.4) | 31 (41.5)  25 (30.5)  2 (2.4)  17 (20.7)  -  4 (4.9) | **0.038** |
| Induction immunotherapy | Alemtuzumab*  IL2 receptor blocker  None/Unknown | 636 (69.3)  103 (11.2)  179 (19.5) | 48 (58.5)  26 (31.7)  8 (9.7) | **0.048** |
| Transplant number | 1^st^  ≥2^nd^ | 806 (57.6)  114 (12.4) | 73 (89.0)  9 (11.0) | 0.71 |
| Diabetes | No  Yes | 592 (64.3)  328 (35.7) | 61 (74.4)  21 (25.6) | 0.07 |
| Vaccine type | BNT1262b2  ChAdOx1 | 490 (53.3)  430 (46.7) | 31 (37.8)  51 (62.2) | **0.0007** |
| Time between vaccinations | Days (median) | 74 (66-77) | 63 (63-64) | **<0.0001** |
| Time of serological test post-boost | Days (median) | 31 (27-35) | 31 (29-31) | 0.90 |

### **Table S3. Characteristics of infection naïve patients undergoing assessment of serological and cellular responses by vaccine type**

| Characteristic | | BNT162b2  N=40 (%) | ChAdOx1  N=39 (%) | p value |
| --- | --- | --- | --- | --- |
| Gender | Male  Female | 30 (75.0)  10 (25.0) | 25 (64.1)  14 (35.9) | 0.30 |
| Age | Years (Median) | 57 (46-64) | 50 (39-56) | **0.016** |
| Ethnicity | Caucasian  Black  Indoasian  Other | 18 (45.0)  2 (5.0)  14 (35.0)  6 (15.0) | 19 (48.7)  6 (15.4)  8 (20.5)  6 (15.4) | 0.30 |
| Cause of ESKD | Glomerulonephritis*  Other | 10 (25.0)  30 (75.0) | 16 (41.0)  23 (59.0) | **0.026** |
| Vaccinated ≤1 year post transplant | Yes  No | 3 (7.5)  37 (92.5) | 25 (64.1)  14 (35.9) | **<0.0001** |
| Immunosuppression at diagnosis | CNI monotherapy*  Other | 21 (52.5)  19 (47.5) | 10 (25.6)  29 (74.4) | **0.015** |
| Induction immunotherapy | Alemtuzumab  Other | 29 (72.5)  11 (27.5) | 16 (41.0)  23 (59.0) | **0.0005** |
| Transplant number | 1^st^  ≥2^nd^ | 36 (90.0)  4 (10.0) | 33 (84.6)  6 (15.4) | 0.47 |
| Diabetes | No  Yes | 23 (57.5)  17 (42.5) | 32 (82.1)  7 (17.9) | **0.018** |

### **Figure S3. Correlation between anti-S (BAU/ml) and T-cell responses (SFU/10^6^ PMBCs)**

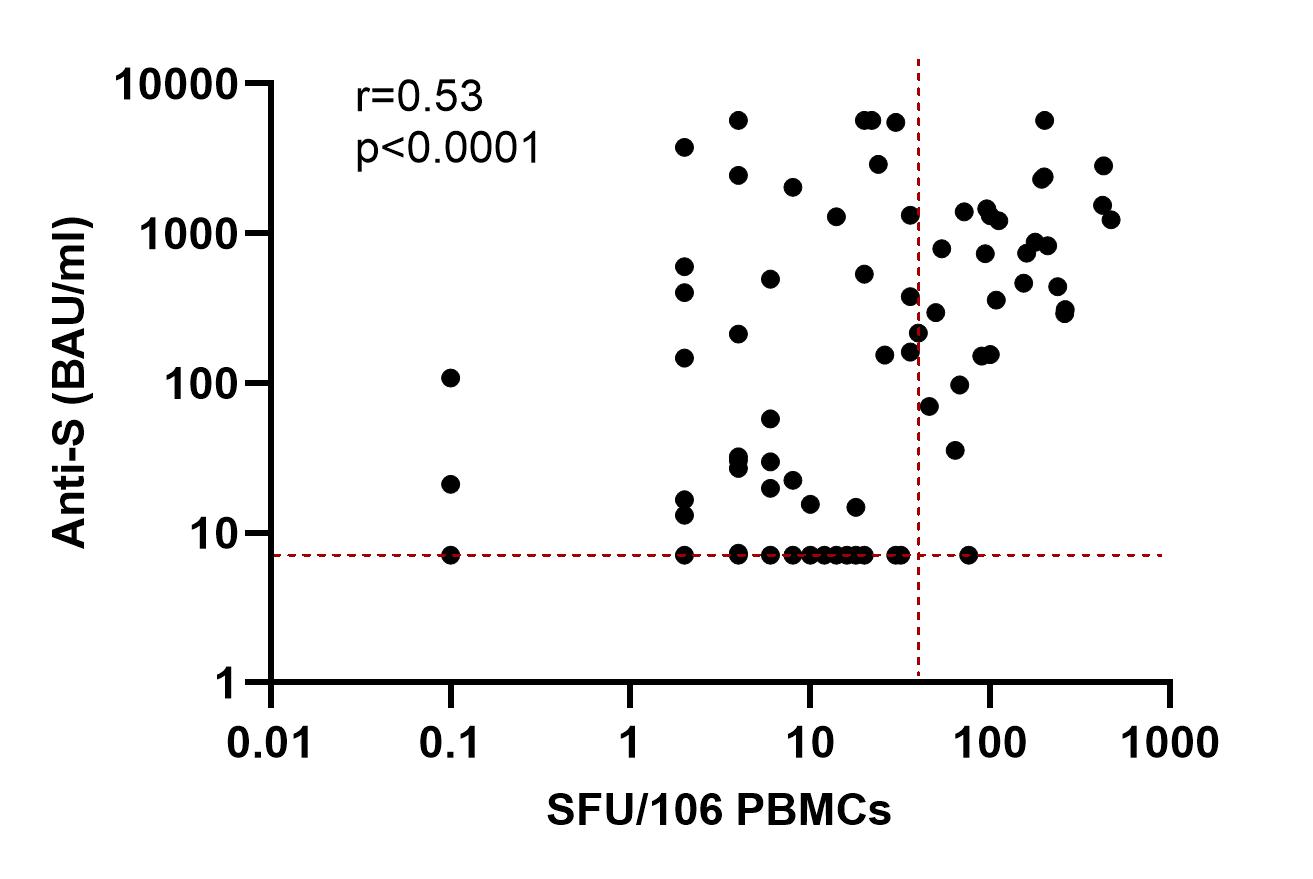

### **Table S4. Characteristics associated with lack of detectable immunological (serological and T-cell) response in transplant patients**

| **Variable** | **Reference Group** | **Univariable** | | **Multivariable** | |
| --- | --- | --- | --- | --- | --- |
|  |  | **OR (95% CI)** | **p value** | **OR (95% CI)** | **p value** |
| **Time of vaccine post-transplant** | <1 year | 0.15 (0.05-0.46) | 0.0009 | 0.14 (0.04-0.57) | 0.006 |
| **Transplant induction agent** | Alemtuzumab | 4.88 (1.87-13.79) | 0.0017 | - | - |
| **Maintenance Immunosuppression** | CNI monotherapy | 15.8 (5.1-49.1) | <0.0001 | 16.5 (4.7-58.0) | <0.0001 |
| **Vaccine** | ChAdOx1 | 0.21 (0.08-0.54) | 0.0013 | - | - |
| **Time to testing post-booster vaccine** | Days | 0.96 (0.90-1.03) | 0.035 | - | - |
